# Cognitive training reshapes functional lateralization of fronto-parietal network in patients with vascular cognitive impairment no dementia

**DOI:** 10.1101/2024.04.18.24306005

**Authors:** Xinhu Jin, Yi Xing, Baihan Lyu, Xiuyi Wang, Yi Tang, Yi Du

**Author notes:** These authors contributed equally to this work (Xinhu Jin, Yi Xing). Corresponding author Yi Tang, Department of Neurology & Innovation Center for Neurological Disorders, Xuanwu Hospital, Capital Medical University, 45 Changchun Street, Beijing 100053, China, (Y.T.), Yi Du, CAS Key Laboratory of Behavioral Science, Institute of Psychology, Chinese Academy of Sciences, 16 Lincui Street, Beijing 100101, China, (Y.D.).

## Abstract

**Background and Objectives:** Vascular cognitive impairment no dementia represents cognitive deficits due to vascular causes but falls short of a dementia diagnosis. Cognitive training has emerged as a safe and effective intervention for vascular cognitive impairment no dementia, though its underlying mechanisms remain obscure. This study investigates how subcortical vascular cognitive impairment no dementia and computerized cognitive training affect a brain fundamental organization feature named functional lateralization.

**Methods:** In a randomized, active-controlled trial, patients with subcortical vascular cognitive impairment no dementia were divided into training and control groups, and underwent neuropsychological assessments and resting-state functional magnetic resonance imaging at baseline, end of 7-week intervention, and 6-month follow-up. Additionally, a healthy older group provided baseline data. Two types of functional lateralization indices (LIs) were defined based on resting-state functional connectivity: LI of intrahemispheric functional connectivity (LI_intra) which represents the left-right difference of functional connectivity strength within the same hemisphere, and LI of interhemispheric heterotopic functional connectivity (LI_he) which represents the left-right difference of functional connectivity strength across bilateral hemispheres.

**Results:** Initially, patients (28 in training group and 28 in control group) showed a fronto-parietal network lateralization pattern akin to healthy older adults (N = 26). However, enhanced right-lateralized LI_he was associated with better memory solely in healthy adults. After the intervention, only the training group exhibited a reduction in LI_he in the fronto-parietal network, indicating a lateralization shift towards bilateral network connectivity. This shift, especially towards leftward lateralization, was linked with improved executive and memory functions in the training group. Notably, these changes disappeared at the 6-month follow-up.

**Discussion:** These findings suggest that subcortical vascular cognitive impairment no dementia modifies the relationship between fronto-parietal network lateralization and cognitive function, rather than altering the lateralization pattern itself. Through hemispherical reorganizing and balancing of the fronto-parietal network, short-term computerized cognitive training facilitates executive and memory functions by leveraging functional compensation by reorganization. This study illuminates the neural plasticity induced by cognitive training in vascular cognitive impairment no dementia, highlighting its potential to transform cognitive outcomes by tapping into the brain’s capacity for reorganization and adaptation.

**Trial registration:** The trial was registered under ClinicalTrials.gov (NCT02640716) and conducted under both CONSORT statement and CONSORT statement for nonpharmacological interventions.

## Introduction

The majority of elderly people experience cognitive decline during aging, with some developing pathological cognitive impairments like dementia ^1,2^. Mild cognitive impairment (MCI) is the critical stage for both preventing and treating dementia. In China, vascular cognitive impairment no dementia (VCIND) is the predominant form of MCI, constituting 42% of cases ^3^. VCIND is characterized by cognitive deficits linked to vascular causes, falling short of dementia diagnosis criteria ^4^. Early intervention of VCIND holds the potential to delay or reverse cognitive impairment ^5^. Subcortical VCIND, a common subtype caused by subcortical ischemic small vessel disease, presents a uniform profile, making it an ideal subject for intervention trials ^6^. However, no intervention for VCIND has been approved. Cognitive training, a structured intervention targeting specific cognitive functions, has been shown to improve the general cognitive function in subcortical VCIND patients ^6^, but the underlying brain mechanisms remain underexplored.

This study investigated the mechanisms through the lens of brain functional lateralization, a fundamental organization feature gauging the differences between the left and right hemispheres ^7,8^. In healthy young adults, language processing is typically specialized in the left hemisphere, while visuospatial tasks show a preference for the right hemisphere ^7,8^. However, in older adults, there is a notable reduction in lateralization and increased activation in the homologous regions of the contralateral hemisphere during tasks ^9,10^. Theories like the Hemispheric Asymmetry Reduction in Older Adults (HAROLD) and the Scaffolding Theory of Aging and Cognition (STAC) models interpret the lateralization reduction in high-performing seniors as a compensatory mechanism ^11,12^. Notably, the effectiveness of this compensation depends on the complementary roles of additionally engaged regions. In some cases, maintaining or enhancing lateralization correlates with improved performance in older adults ^13,14^. Hence, linking functional lateralization to behavioral performance is essential to differentiate compensation from other mechanisms like neural inefficiency, dedifferentiation, or pathology ^15^. In addition, the recruitment of regions in the right hemisphere, which do not typically support language processing, is frequently observed in the recovery from aphasia following a left-hemisphere stroke ^16^. Research on stroke patients has demonstrated a correlation between the severity of behavioral impairment following focal neural damage and the extent of activation and connectivity changes in remote regions ^17,18^. These findings imply that compensation by reorganization in patient populations may not be constrained by the intrinsic functions of recruited regions but rather by structurally or functionally connected connectomics ^19^.

Furthermore, neurodegenerative disorders like MCI and Alzheimer’s disease (AD) can alter brain structural and functional lateralization ^20,21^. Presently, no evidence exists of lateralization changes in subcortical VINCD or the impact of cognitive training on such lateralization. Previous studies have demonstrated that cognitive training improves executive function and episodic memory in healthy seniors and MCI patients ^22,23^, processes closely linked to the fronto-parietal network (FPN) in the brain ^24,25^. A meta-analysis study has unveiled that cognitive training boosts activation in left FPN regions and reduces it in right frontal regions in healthy older adults, without observing changes elsewhere ^26^. This suggests that the functional lateralization of FPN is particularly responsive to cognitive training. Similarly, cognitive training is found to retain the intrinsic FPN lateralization pattern, contrasting with its reduction in a control group of healthy older adults ^27^. In our prior work using the same dataset of the present study, we have uncovered an increased resting-state functional connectivity (rsFC) between the left dorsolateral prefrontal cortex (a FPN region) and medial prefrontal cortex (a default mode network region) in subcortical VCIND patients after a 7-week cognitive intervention, correlating with improvement in Montreal Cognitive Assessment (MoCA) ^6^. In light of executive dysfunction and deficits in episodic memory being prominent characteristics of VCIND ^28,29^, and the pivotal role of FPN in cognitive control and episodic memory ^24,25^, coupled with the demonstrated sensitivity of FPN activity to cognitive training ^6,26,27^, we hypothesized that cognitive training may improve executive function and episodic memory in subcortical VCIND patients through modulation of the functional lateralization in FPN.

This resting-state functional magnetic resonance imaging (rs-fMRI) study aimed to uncover the characteristic FPN lateralization pattern in subcortical VCIND patients compared to healthy older adults, and how multidomain cognitive training enhanced executive function and episodic memory by altering FPN lateralization. We focused on three primary outcome measures: the Trail Making Test (TMT) for executive function ^30^, the Auditory Verbal Learning Test (AVLT) for episodic memory ^31^, and the MoCA for global cognitive function ^32^. We defined two functional lateralization indices (LIs) of FPN based on rsFC ^33–36^: LI of interhemispheric heterotopic FC (LI_he), denoting the left-right difference of FC strength across the bilateral hemispheres, and LI of intrahemispheric FC (LI_intra), representing the left-right difference of FC strength within the same hemisphere. Our hypotheses were twofold. First, compared to healthy older controls, subcortical VCIND patients might exhibit a distinct FPN lateralization pattern or varying correlations between FPN lateralization and task performance that rely on FPN functioning. Second, we aimed to test two alternative hypotheses regarding the effect of cognitive training on FPN lateralization. According to the compensation hypothesis, cognitive training would result in a reduction in FPN lateralization, correlating with better performance after training. In contrast, the complementary hypothesis predicts that the effect of cognitive training on FPN lateralization is contingent upon whether the opposite regions are complementary to task performance. In the case of the TMT task, which relies on visual search and working memory presumed to be specialized within the right FPN ^37^, diminished FPN lateralization may not compensate for performance. Conversely, for the AVLT task, which taps both the control of verbal processes that tend to be left-lateralized in the prefrontal cortex and memory retrieval which often shows right-lateralized prefrontal activation ^38^, FPN with bilaterally symmetrical interaction would be beneficial for performance.

## Materials and methods

### Study design and registrations

The present randomized, active-controlled clinical trial was conducted under both the Consolidated Standards of Reporting Trials (CONSORT) statement and the CONSORT statement for nonpharmacological interventions. Participants were recruited from three centers: Xuanwu Hospital; Fu Xing Hospital and Beijing Friendship Hospital, Capital Medical University. All participants provided written informed consent. Ethical approval was obtained from the Ethics Committee of Xuanwu Hospital, Capital Medical University. The trial was registered under ClinicalTrials.gov (NCT02640716) and its protocol has been published previously ^5^.

### Participants

Participants were enrolled from December 22, 2015 through November 7, 2016. The last follow-up measurements were obtained on May 8, 2017. A total of 212 individuals from the neurology and geriatric clinics were included and assessed for eligibility. The diagnosis of subcortical VCIND was based on evidence of both cognitive impairment without dementia and small vessel ischemic disease. All patients, who met the inclusion criteria, were diagnosed by a consensus panel including three senior neurologists. For specific inclusion criteria, see the corresponding section in the previously published articles that utilized the same groups of participants ^5,6^. We excluded participants who exhibited any condition that would preclude completion of neuropsychological testing or disorders other than subcortical VCIND that would affect cognition. Finally, sixty right-handed patients with subcortical VCIND were recruited on fulfillment of the inclusion and exclusion criteria. In addition, we recruited thirty healthy older adults as the healthy group (HG), who showed no significant group differences in age and education compared to subcortical VCIND patients. They did not have any cognitive impairment complaints and attained a score of zero on the clinical dementia rating. All MRI and behavioral data of healthy older adults were acquired only at the baseline 0.

All sixty subcortical VCIND patients were randomly assigned to the training group (TG) or the control group (CG), with thirty in each group. The intervention began directly after randomization. Patients in the TG received a computerized, adaptive, multidomain training program for seven weeks, whereas patients in the CG received fixed processing speed and attention tasks set to a primary difficulty level. All patients completed the tasks at home and were supervised by an independent neurologist over the Internet (www.66nao.com) to guarantee the fulfillment of training. All MRI and behavioral data of VCIND patients were acquired at baseline 0, end of intervention (weeks 7), and six months after randomization (months 6). Finally, a total of 82 participants (28 in TG, 28 in CG, and 26 in HG) achieved the baseline data acquisition with qualified and available data. A total of 49 participants (24 in TG and 25 in CG) finished the 7-week cognitive training intervention trial with qualified and available data. A total of 35 participants (14 in TG and 21 in CG) completed the 6-month follow-up with qualified and available data. The flow of participants through the study is shown in Fig. 1. For more specific details, see the corresponding section in the previously published articles ^5,6^.

**Fig.1.**
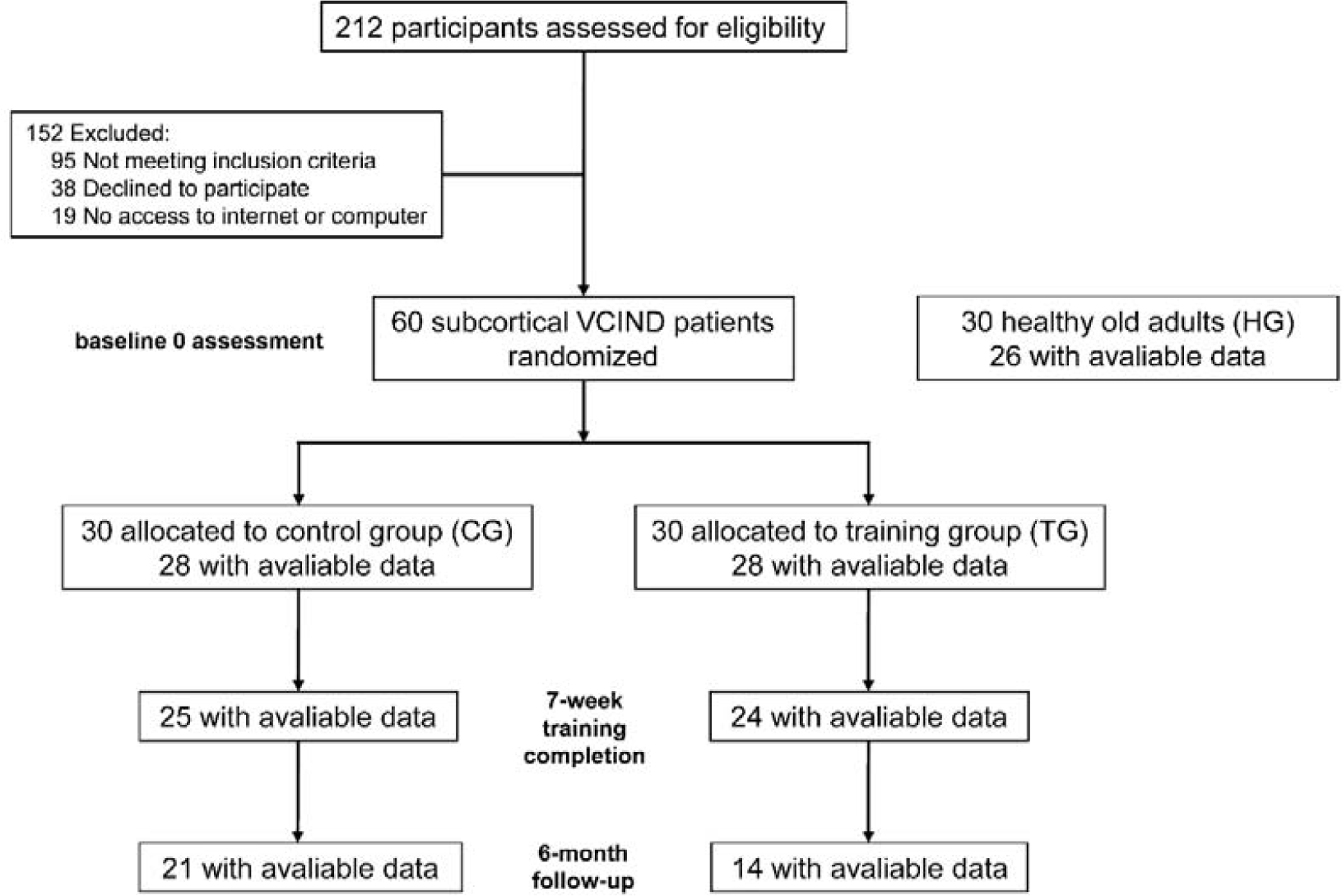
The flowchart for participants recruitment, intervention and assessment in this study.

### Neuropsychological Assessments

Given the neuropsychological profile of VCIND, characterized by prominent impairments in executive function and episodic memory ^28,29^, and our research focus on cognitive functions closely related to the FPN, we adopted three tests as primary outcomes: the TMT for executive function ^30^, the WHO-UCLA AVLT for episodic memory ^31^, and the MoCA for global cognitive function ^32^. We also assessed secondary outcomes utilizing the Boston Naming Test (BNT) and Digit Span. In our previous work, 7 weeks of cognitive training significantly improved performance only in MoCA and BNT but not in other measures ^6^.

### MRI Data Acquisition and Processing

The MRI data were collected using a 3T Siemens scanner (Magnetom Skyra). Resting-state fMRI was measured using echo-planar imaging (EPI) series while participants were instructed to rest still and quietly (repetition time (TR) = 2000 ms, echo time (TE) = 30 ms, field of view (FOV) = 224 mm, flip angle (FA) = 77°, voxel size = 3.5 × 3.5 × 3.5 mm^3^, 35 slices). A high-resolution T1-weighted anatomical image was obtained using a sagittal 3D magnetization-prepared rapid gradient echo (MP-RAGE) sequence (TR = 2100 ms, TE = 2.26 ms, FOV = 256 mm, FA = 12°, slice thickness = 1 mm).

Preprocessing was performed using fMRIPrep 20.2.5 ^39^. Rs-fMRI data were preprocessed with slice-timing correction, motion correction, distortion correction, co-registration to structural data, normalization to Montreal Neurological Institute (MNI) space, and projection to cortical surface. Built with Nipype 1.7.0 ^40^, the eXtensible Connectivity Pipeline (XCP-D) ^41^ was used to post-process the outputs of fMRIPrep. For each CIFTI run found per participant, all data were subjected to demeaning, detrending, and nuisance regression. Volumes with framewise displacement (FD) greater than 0.5 mm were flagged as outliers and excluded from nuisance regression ^42,43^. Specifically, images were de-noised using a 36-parameter confound regression model that has been shown to minimize associations with motion artifact while retaining signals of interest in distinct sub-networks ^44^. This model included the six framewise estimates of motion, the mean signal extracted from eroded white matter and cerebrospinal fluid compartments, the mean signal extracted from the entire brain, the derivatives of each of these nine parameters, and quadratic terms of each of the nine parameters and their derivatives. Next, the residual time series from this regression were band-pass filtered to retain signals within the 0.01-0.08 Hz frequency band. By adopting “scrubbing” for each of these data points (head radius = 50 mm for computing FD, FD threshold = 0.5 mm for censoring), we finally excluded two participants in HG and two in CG with more than 20% data above the high motion cutoff (FD > 0.5). Moreover, two participants in HG and two in TG for lacking complete MRI or behavioral data were also excluded. Therefore, eighty-two right-handed participants (28 in TG, 28 in CG, and 26 in HG) at baseline 0 were included in the subsequent analysis.

### Calculation of LI and LI difference of FPN

To quantify the functional connectivity at the surface level, a multimodal parcellation of 360 areas (180 per hemisphere) was applied, which is suitable for studying asymmetry across homologous brain parcels ^45^. The rsFC between two parcels was computed as the Pearson correlation (*r*) of the two BOLD time series. Fisher’s r-to-z transformation was further applied to the original correlation coefficient *r* values to improve the normality.

Based on the whole-brain rsFC (z), we obtained interhemispheric heterotopic FC and intrahemispheric FC. The former represents the FC between two cortical parcels across different hemispheres, except the homotopic pairs, while the latter refers to the FC between two cortical parcels within the same hemisphere. For a specific parcel, the heterotopic (he) was defined as the sum of heterotopic FCs between this parcel and all the others in the opposite hemisphere except the homotopic one, while the intrahemispheric (intra) was defined as the sum of intrahemispheric FCs between this parcel and all the others within the same hemisphere. On these bases, for a pair of homotopic parcels, we further defined two different forms of functional lateralization (Fig. 2A), calculated as LI_he (equation 1) and LI_intra (equation 2). These two LI measures have been validated by previous studies ^34–36^.

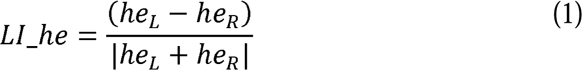

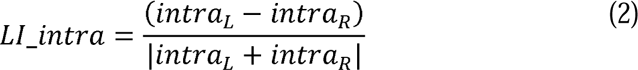

**Fig.2.**
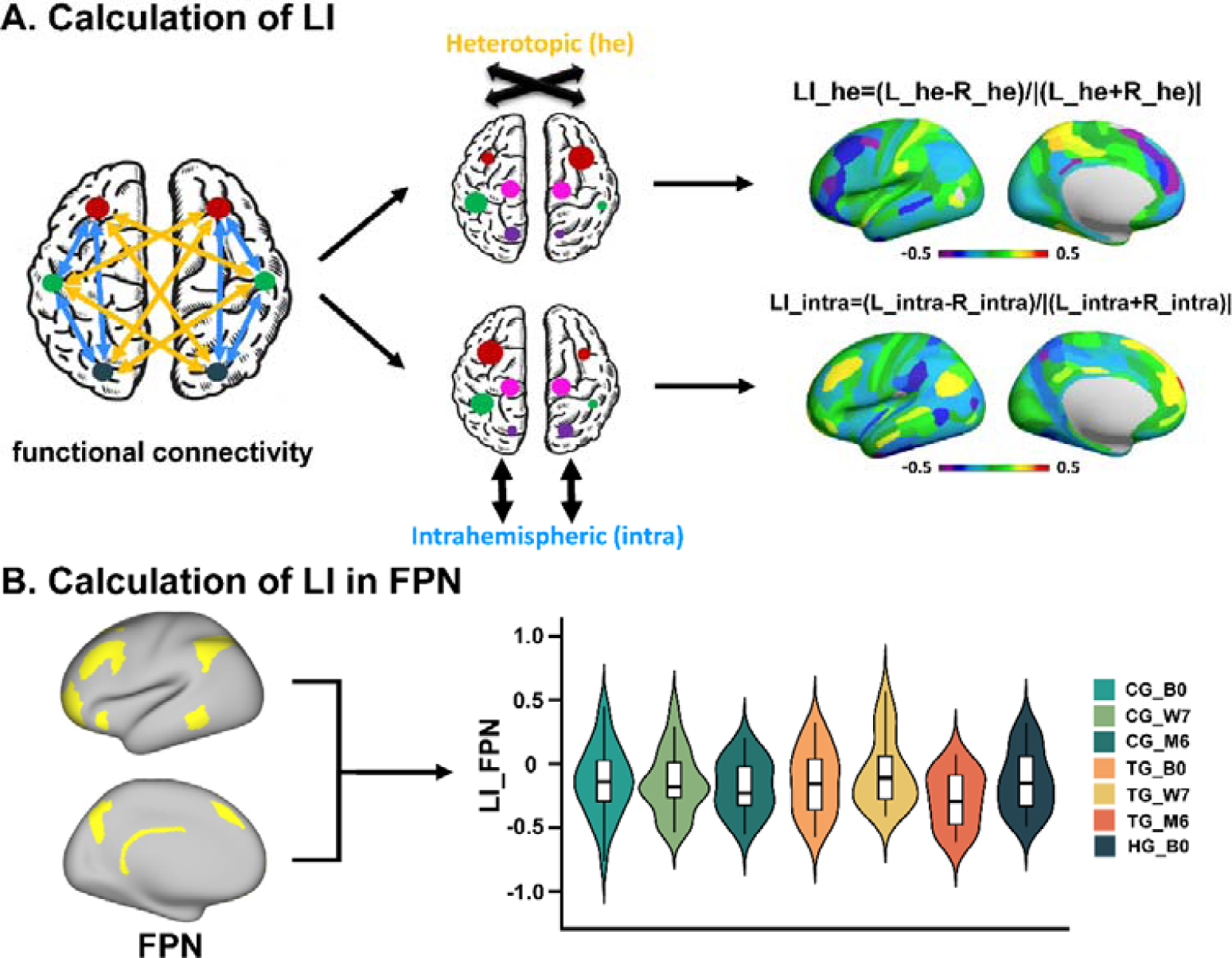
Workflow of analyses. (**A**) Calculation of LI. We first defined two different types of functional connectivity (FC) among the whole brain, named interhemispheric heterotopic FC (yellow) and intrahemispheric FC (blue). For a specific parcel, the heterotopic (he) was defined as the sum of heterotopic FCs between this parcel and all the others in the opposite hemisphere except the homotopic one, whereas the intrahemispheric (intra) was defined as the sum of intrahemispheric FCs between this parcel and all the others within the same hemisphere. The functional lateralization between each homotopic pair of parcels was quantified by a commonly used laterality index (LI): LI = (L-R)/|(L+R)|. Finally, every participant would have two different LI maps of he and intra. Larger positive values of LI_he and LI_intra imply stronger across-hemispheric interactions or within-hemispheric interactions in left-hemispheric parcels, respectively, whereas larger negative values indicate stronger interactions in right-hemispheric parcels. (**B**) Calculation of LI in FPN. Based on Cole-Anticevic Brain-wide Network Partition version 1.0 (CAB-NP v1.0), we averaged LIs of homotopic parcels both belonging to the frontoparietal network (FPN) and acquired the LI of FPN in every participant at each time point. CG/TG/HG_B0, control/training/healthy group at baseline 0; CG/TG_W7, control/training group at the end of the 7-week intervention; CG/TG_M6, control/training group at the 6-month follow-up.

According to the Cole-Anticevic Brain-wide Network Partition version 1.0 (CAB-NP v1.0) ^46^, all cortical parcels were mapped into twelve networks. Since some homotopic pairs of parcels were labeled to different networks, we only chose the homotopic pairs of parcels whose left and right parcels both belong to the FPN to calculate the LI of FPN. After averaging the LIs across these homotopic pairs of FPN parcels, we acquired the LIs of FPN for each participant. Larger positive values of LI_he and LI_intra of FPN imply stronger bilateral across-hemisphere interactions or stronger ipsilateral within-hemisphere interactions in left-hemispheric FPN nodes, whereas larger negative values indicate stronger interactions in right-hemispheric FPN nodes. Then we obtained the LI difference of FPN by subtracting the LI of FPN at baseline 0 from it at the other two latter time points (weeks 7 and months 6) for participants who had completed the corresponding MRI data collection. By doing so, LIs of FPN (CG and TG: baseline 0, weeks 7, months 6; HG: baseline 0) and LI differences of FPN (CG and TG: weeks 7 - baseline 0, months 6 – baseline 0) were obtained (Fig. 2B). All calculations above were performed on both LI_he and LI_intra of FPN.

### Statistical Analysis

The differences in demographic and neuropsychological data were tested by independent two-sample t-tests between two subcortical VCIND groups and one-way analyses of variance (ANOVAs) as well as one-way analyses of covariance (ANCOVAs) among the healthy group and two subcortical VCIND groups. Sex differences were tested by Chi-square tests. To highlight the differences in LI_he and LI_intra between the subcortical VCIND patients and healthy adults, we presented the violin maps of LIs of FPN in HG, CG, and TG at baseline 0, respectively. One-way ANCOVAs were adopted to compare the group differences in LIs of FPN. Next, we performed separate one-sample t-tests on LIs to identify whether the FPN was lateralized (leftward/rightward) or symmetric in each group. In addition, partial correlations between LIs of FPN and neuropsychological scores were performed in three groups respectively, considering relevant confounding variables including age, sex, education, mean framewise displacement (mFD), and mean of rsFCs between any two cortical parcels in the whole brain (mFC) ^47–51^.

We then explored the effect of cognitive training on LIs of FPN in subcortical VCIND patients, adopting linear mixed-effect models nested within individuals. We analyzed the LI_he/LI_intra differences of FPN from baseline 0 to end of intervention (weeks 7) and from baseline 0 to 6-month follow-up (months 6). Group (control/training), time (weeks 7/months 6), and group-by-time were included as fixed effects, and participant was assigned as the repeated variable. Age, sex, education, mFD, and mFC were treated as covariates. Finally, partial correlations between LI differences of FPN and neuropsychological scores were performed, regressing the same confounding variables. All the results in our analyses were considered to be significant with a value below 0.05 after FDR correction or Games-Howell correction (with heterogeneity of variance).

## Results

### Baseline neuropsychological performance

Baseline characteristics and neuropsychological assessments of participants are shown in **Table 1**. We found no group difference in age, sex, education, and neuropsychological testing scores between the two subcortical VCIND groups. While among HG and VCIND groups, age and education were matched except for sex (*χ*^2^ = 11.084, *p* = 0.004). Using sex as the covariate, significant group differences were discovered between HG and VCIND groups in all neuropsychological scores (all *F/Welch F* > 4.338, η*_p_*^2^ > 0.100, *p* < 0.05) except TMT B-A (*F*_(2, 79)_ = 1.333, *p* = 0.270) and BNT (*F*_(2, 79)_ = 2.452, *p* = 0.093), indicating impaired executive function (TMT-B), episodic memory, working memory and general cognition in subcortical VICND patients than healthy older adults.

**Table 1.** The group mean ± standard deviation values and statistics of demographic and behavioral data at baseline 0 in each group.

| | Control group<br>(n=28) | Training group<br>(n=28) | $t/\chi^2$ ($p$) | Healthy group<br>(n=26) | $F/\chi^2/Welch F$ ($p$) |
| --- | --- | --- | --- | --- | --- |
| Age (year) | 65.32±6.62 | 65.68±8.02 | -0.182<sup>a</sup> (0.856) | 62.65±6.11 | 1.493<sup>c</sup> (0.231) |
| Sex (F/M) | 8/20 | 12/16 | 1.244<sup>b</sup> (0.265) | 19/7 | 11.084<sup>b</sup> (0.004) |
| Education (year) | 10.11±2.82 | 10.61±3.45 | -0.594<sup>a</sup> (0.555) | 11.42±2.87 | 1.261<sup>c</sup> (0.289) |
| MoCA (score) | 21.25±3.76 | 21.79±3.88 | -0.525<sup>a</sup> (0.602) | 26.69±1.76 | 21.991<sup>d</sup> (<0.001) |
| TMT A (second) | 76.92±33.50 | 83.60±43.22 | -0.646<sup>a</sup> (0.521) | 42.51±12.84 | 12.206<sup>d</sup> (<0.001) |
| TMT B (second) | 151.31±78.63 | 161.21±86.56 | -0.448<sup>a</sup> (0.656) | 83.33±52.35 | 9.513<sup>d</sup> (<0.001) |
| TMT B-A (second) | 74.40±64.99 | 77.61±56.86 | -0.197<sup>a</sup> (0.845) | 40.82±46.75 | 1.333<sup>e</sup> (0.270) |
| AVLT_Immediate recall (number) | 19.54±6.64 | 22.79±6.01 | -1.920<sup>a</sup> (0.060) | 29.12±4.76 | 19.561<sup>d</sup> (<0.001) |
| AVLT_Delayed recall (number) | 6.50±3.16 | 7.29±2.77 | -0.989<sup>a</sup> (0.327) | 11.12±2.72 | 13.966<sup>e</sup> (<0.001) |
| AVLT_Recognition (number) | 10.07±2.84 | 11.07±2.75 | -1.339<sup>a</sup> (0.186) | 13.12±1.84 | 11.155<sup>d</sup> (<0.001) |
| Digit span forward | 7.18±1.12 | 7.25±1.53 | -0.199<sup>a</sup> (0.843) | 8.46±0.86 | 6.748<sup>e</sup> (0.002) |
| Digit span backward | 4.25±1.01 | 4.25±1.51 | 0.000<sup>a</sup> (1.000) | 5.23±1.48 | 4.338<sup>e</sup> (0.016) |
| BNT | 23.61±3.52 | 22.00±3.83 | 1.635<sup>a</sup> (0.108) | 24.85±5.56 | 2.452<sup>e</sup> (0.093) |

MoCA, Montreal Cognitive Assessment; TMT, Trail Making Test; AVLT, WHO-UCLA Auditory Verbal Learning Test; BNT, Boston Naming Test.
<sup>a</sup> Independent two-sample t-test.
<sup>b</sup> Chi-square test.
<sup>c</sup> One-way analysis of variance with $F$ value.
<sup>d</sup> One-way analysis of covariance with $Welch F$ value.
<sup>e</sup> One-way analysis of covariance with $F$ value.

### FPN lateralization differences between healthy older adults and subcortical VCIND patients

To reveal how subcortical VCIND changes the lateralization pattern of FPN, we first explored the functional lateralization pattern of FPN in three groups at baseline 0. We hypothesized that VCIND patients may exhibit disparate FPN lateralization patterns or changed relationships between FPN lateralization and behavioral performances in executive and memory tasks. Since no group difference existed on mFD (*F*_(2, 79)_ = 0.670, *p* = 0.515) and mFC (*F*_(2, 79)_ = 0.471, *p* = 0.626) among the three groups, one-way ANCOVAs only controlling for sex revealed no significant group difference on both LI_he (*F*_(2, 79)_ = 0.003, *p* = 0.997) and LI_intra (*F*_(2, 79)_ = 0.383, *p* = 0.683), indicating similar functional lateralization patterns of FPN in healthy older adults and subcortical VCIND patients. One-sample t-tests found significantly right-lateralized LI_he (all *t* < −2.615, all Cohen’s *d* < −0.494, FDR-corrected *p* = 0.014) but bilateral LI_intra in all three groups (**Fig. 3A**). We then asked whether and how functional lateralization of FPN was related to neuropsychological performances. By regressing age, sex, education, mFD, and mFC, we conducted partial correlations between LIs of FPN and behavioral scores in three groups, respectively. As shown in **Fig. 3B**, significant negative correlations between LI_he and AVLT scores were only discovered in HG (immediate recall: *r* = −0.456, FDR-corrected *p* = 0.038; delayed recall: *r* = −0.511, FDR-corrected *p* = 0.038; recognition: *r* = −0.467, FDR-corrected *p* = 0.038), suggesting that the more right-lateralized LI_he in FPN, the better memory performance in healthy older adults. For LI_intra, more right-lateralized FPN was only negatively correlated with better AVLT recognition in HG (*r* = −0.458, uncorrected *p* = 0.037). However, there was no other significant correlation between LIs of FPN and behaviors in these three groups.

**Fig.3.**
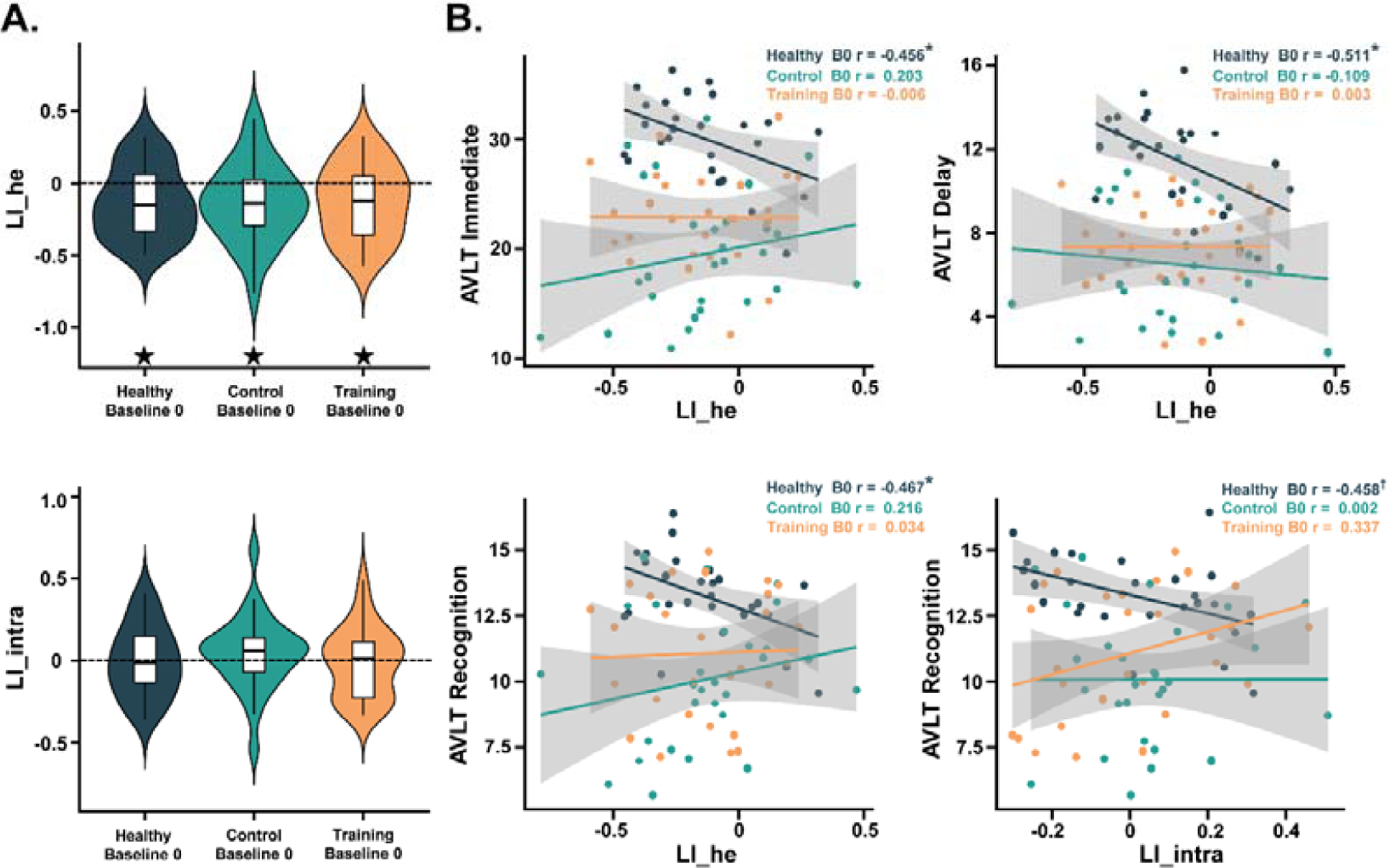
FPN lateralization patterns and their relationships with behaviors in three groups at baseline 0. (**A**) LI_he and LI_intra of FPN in HG, CG, and TG at baseline 0. ^11^ FDR-corrected *p* < 0.05 by one-sample t-tests. All dashed lines indicate zero (no functional lateralization). (**B**) Partial correlations between LIs of FPN and AVLT performances in HG, CG, and TG at baseline 0, after controlling for age, sex, education, mean framewise displacement (mFD) and mean global functional connectivity (mFC). * FDR-corrected p < 0.05, † uncorrected p < 0.05.

### Cognitive training effect on FPN lateralization in subcortical VCIND patients

To further uncover the effect of cognitive training on functional lateralization of FPN in subcortical VCIND patients, we compared LI_he and LI_intra of FPN in TG and CG at three time points (**Fig. 4A**). We expected to observe a training-induced reduction in the lateralization of FPN which correlated with better performances regardless of tasks, or task-specific correlations between FPN lateralization changes after cognitive training and task performances, depending on whether the task relies on bilateral recruitment. The linear mixed-effect models revealed no significant main effect (0.034 < *F* < 3.008, *p* > 0.05), but a significant group × time interaction for the LI_he difference of FPN after the cognitive intervention (*F*_(1, 34.90)_ = 4.571, *η_p_*^2^ = 0.120, *p* = 0.040) in **Fig. 4B** (weeks 7 - baseline 0: 24 in CG and 23 in TG; months 6 - baseline 0: 21 in CG and 14 in TG). Moreover, one-sample t-tests detected significant right-lateralized LI_he (all *t* < −2.615, all Cohen’s *d* < −0.494, FDR-corrected *p* < 0.05) at all three time points in both groups except in TG at the end of intervention (weeks 7, *t* = −1.194, *p* = 0.245). In other words, there was a significant lateralization change from right-lateralization to bilateralization in FPN after the 7-week intervention in TG. However, this change did not persist at the 6-month follow-up in this group, moving to a more right-lateralized pattern instead. While for LI_intra of FPN, no significant lateralization was detected in both groups at all three time points (−1.526 < *t* < 1.041, *p* > 0.05).

**Fig.4.**
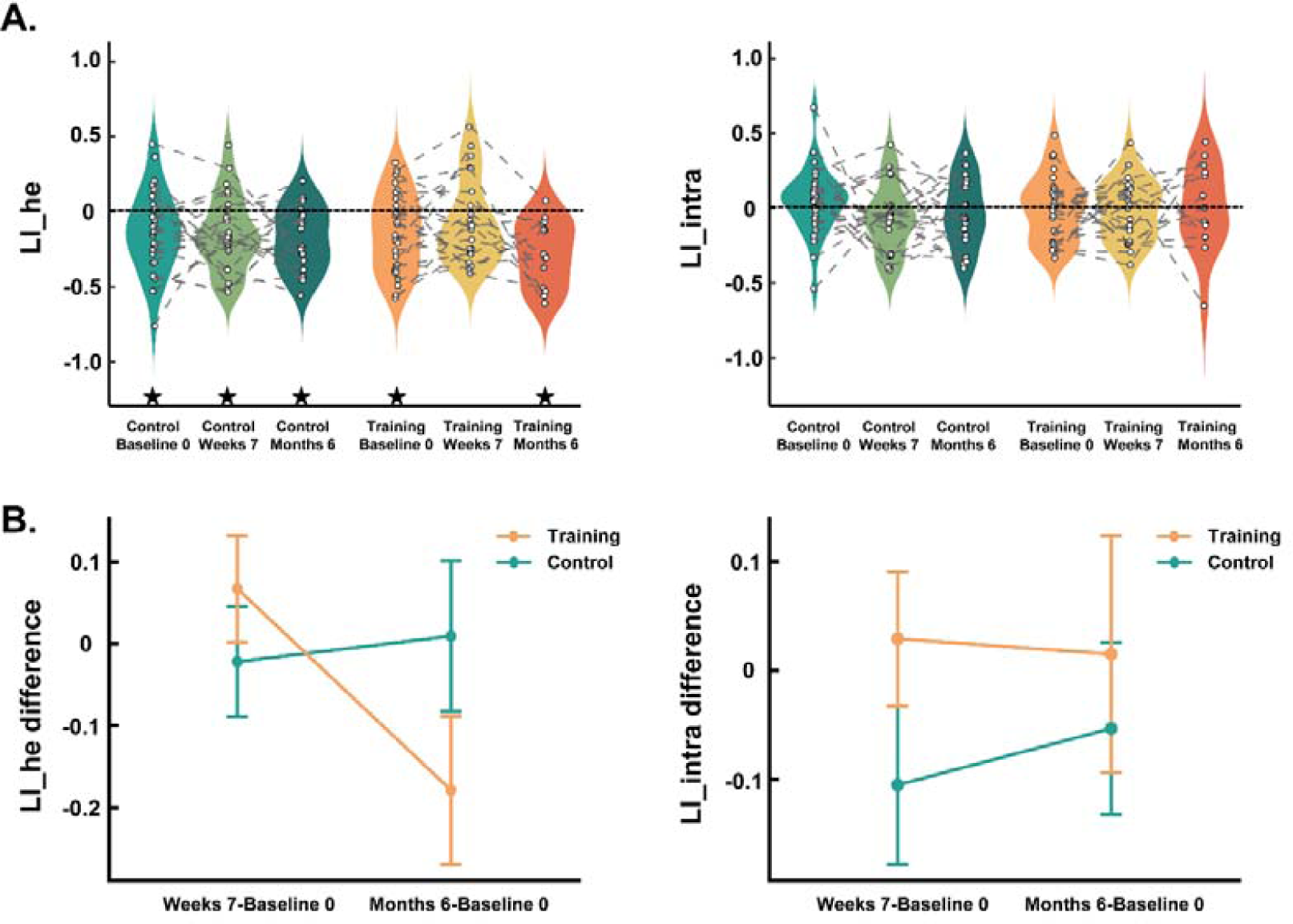
LIs and LI differences of FPN in patients with subcortical VCIND. (**A**) LI_he and LI_intra of FPN were compared among CG and TG at three time points. ^11^ FDR-corrected *p* < 0.05 by one-sample t-tests. All dashed lines indicate zero (no functional lateralization). (**B**) The group (CG/TG) × time (weeks 7 - baseline 0/months 6 - baseline 0) interactions in LI_he/LI_intra difference of FPN.

As for the training effect on neuropsychological assessments, the linear mixed-effect model including age, sex, and education as covariates revealed a significant main effect of time (*F*_(2, 82.41)_ = 15.715, *p* < 0.001) and a significant group (CG/TG) × time (baseline 0/weeks 7/months 6) interaction for the MoCA score (*F*_(2, 80.79)_ = 7.264, *η_p_* = 0.150, *p* = 0.001, **Fig. 5C**). In TG, MoCA significantly improved after 7 weeks of training compared to baseline 0 (*t*_80.5_ = 5.895, *p* < 0.001). While in CG, MoCA had significantly improved after 6 months (*t_82.3_* = 3.562, *p* = 0.008). The linear mixed-effect model also revealed a significant main effect of time (*F*_(2, 80.88)_ = 4.758, *p* = 0.011) and a significant group (CG/TG) × time (baseline 0/weeks 7/months 6) interaction for the BNT score (*F*_(2, 79.14)_ = 7.687, *η*_*p*_^2^ = 0.160, *p* < 0.001, **Fig. 5D**). Only in TG, BNT significantly improved after 7 weeks (*t*_80.1_ = 4.549, *p* < 0.001) and 6 months (*t_84.2_*= 3.389, *p* = 0.013) compared to baseline 0. However, no significant training effect was observed on the TMT, AVLT, and digit span scores (all *F* < 2.846, *p* > 0.064, **Fig. 5A, 5B**).

**Fig.5.**
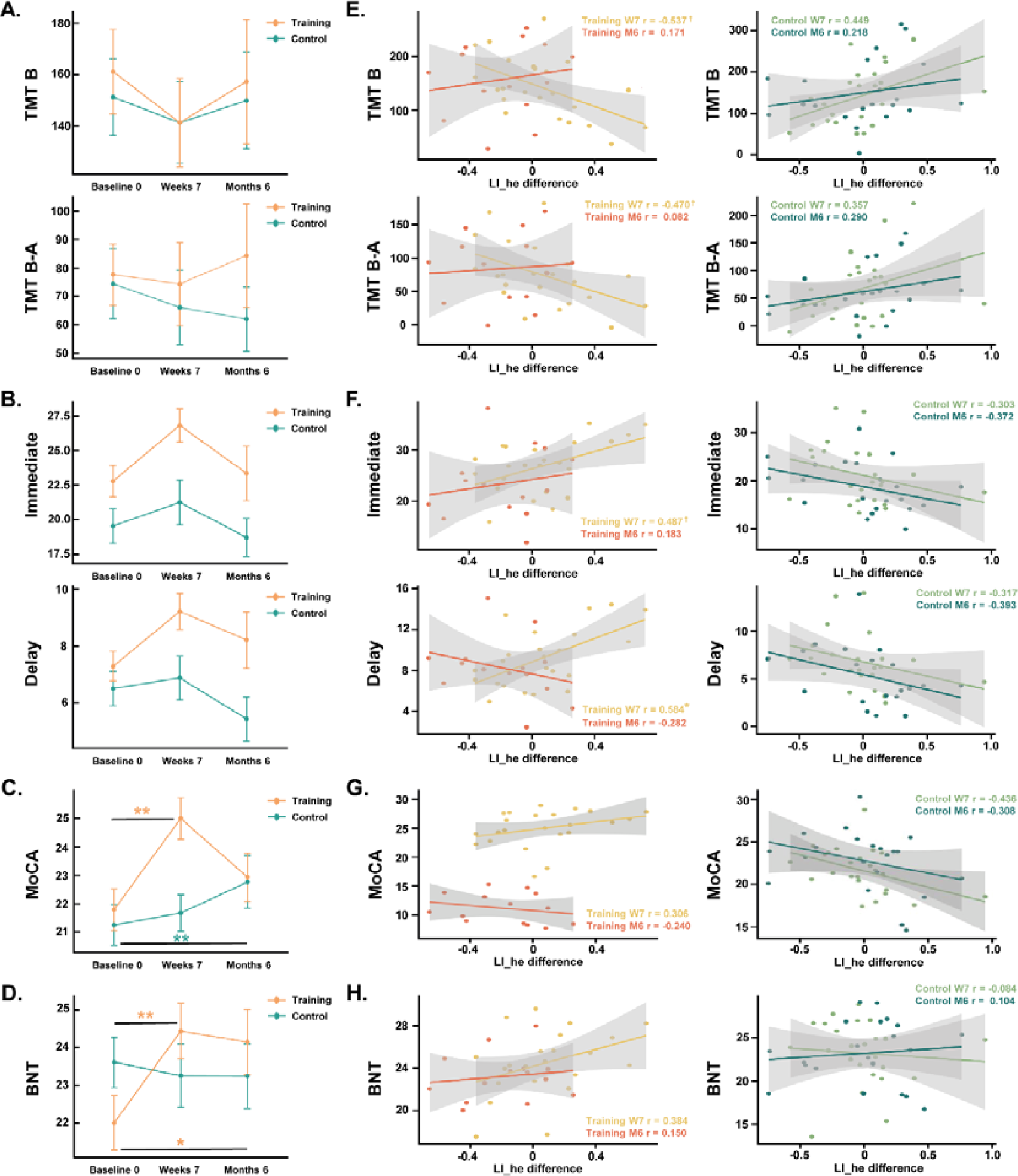
Behavioral performances and correlations between LI differences of FPN and behaviors in subcortical VCIND patients. (**A-D**) Behavioral performances at three time points in TG and CG. ** *p* < 0.01, * *p* < 0.05, marks the significant group×time effect. (**E-H**) Partial correlations between LI_he differences of FPN and TMT (**E**), AVLT (**F**), MoCA (**G**), and BNT (**H**) performances in TG and CG, after controlling for age, sex, education, mFD, and mFC. * FDR-corrected p < 0.05, † uncorrected p < 0.05.

We then explored whether the change of LI_he in FPN at the end of intervention was linked to better cognitive functions in subcortical VCIND patients. Utilizing partial correlations between LI_he differences of FPN and neuropsychological scores, significant correlations were only discovered in TG at the end of the 7-week intervention but not at the 6-month follow-up. In **Fig. 5E**, shorter TMT reaction times were correlated with stronger left-lateralized changes of LI_he in FPN in TG after cognitive training (TMT B: *r* = −0.537, uncorrected *p* = 0.022; TMT B-A: *r* = −0.470, uncorrected *p* = 0.049); that is, the stronger leftwards changes of FPN lateralization after cognitive training, the better executive function of patients. Similarly in the TG, larger AVLT scores were correlated with stronger left-lateralized changes of LI_he in FPN after cognitive training (**Fig. 5F**, immediate recall: *r* = 0.487, uncorrected *p* = 0.040; delayed recall: *r* = 0.584, uncorrected *p* = 0.011, FDR-corrected *p* = 0.033); that is, the greater left-lateralized changes of FPN lateralization after cognitive training, the better memory function of patients. However, there was no significant correlation between LI_he differences and MoCA (**Fig. 5G**, −0.437 < *r* < 0.307, *p* > 0.05), BNT (**Fig. 5H**, −0.085 < *r* < 0.385, *p* > 0.05) as well as digit span (−0.116 < *r* < 0.248, *p* > 0.05) in either group.

## Discussion

From the rarely explored perspective of intrinsic functional lateralization of brain networks, this study provides evidence that short-term multidomain computerized cognitive training alters the functional lateralization pattern of FPN from right-lateralized to bilaterally symmetric in subcortical VCIND patients, which facilitates their executive and memory functions. Prior to training, both subcortical VCIND patients and healthy older adults exhibited comparable right-lateralized interhemispheric heterotopic FC in FPN. However, a stronger right-lateralized LI_he in FPN was associated with better memory performance solely in the healthy group; however, no such relationship was observed in subcortical VCIND patients. These results support our hypothesis that subcortical VCIND alters the relationship between FPN lateralization and behavioral performance rather than the FPN lateralization itself. After 7 weeks of cognitive training, the right lateralization of FPN shifted towards a bilaterally symmetric pattern only in the VCIND training group, but not in the VCIND control group. Moreover, significant correlations were found between differences (from baseline to end of training) in LI_he of FPN and performances on TMT as well as AVLT exclusively within the VCIND training group, indicating that greater leftward changes in LI_he of FPN were linked with better executive and memory functions after intervention. These findings align with compensation theories that short-term cognitive training can compensate for behavior by reorganizing the functional lateralization of FPN in patients with subcortical VCIND, which benefits executive and memory functions reliant on the FPN. Notably, this reorganization does not simply reinstate the original lateralization pattern and its functionality observed in healthy older adults but rather induces beneficial changes by reducing functional lateralization and balancing bilateral functional connectivity of FPN. Our study offers new insights into the potential and mechanism of computerized cognitive training as a therapeutic intervention for cognitive impairments.

### Dysfunction of FPN lateralization in subcortical VCIND patients

FPN plays a central role in cognitive control, which supports a range of cognitive functions, including working memory, encoding and retrieval during episodic memory^25^. In young adults, the interhemispheric connections from the right intraparietal sulcus/frontal eye field to the left counterpart exhibit greater strength than those in the opposite direction, suggesting a right-lateralized interhemispheric connectivity within the FPN ^52^. This FPN lateralization pattern may contribute to cognitive reserve in healthy older adults resistant to neurodegeneration in MCI and AD. Cognitive reserve refers to the phenomenon where older adults who engage in intellectually stimulating environments experience less age-related decline in cognitive abilities and degenerative brain changes attributed to AD ^53^. Previous studies have shown that connectivity in the right FPN is crucial for preserved alertness function among healthy older participants ^54^, and higher levels of cognitive reserve in older adults are associated with increased involvement of the right FPN during visual information processing ^55^. Considering that cognitive reserve develops through repeated activation of norepinephrine with its privileged association with the right FPN underlying arousal, sustained attention, working memory, self-monitoring processes, and novelty, a prominent role for the right FPN in cognitive reserve is proposed ^37^.

Consistent with this idea, this study observed a significant correlation between right-lateralized LI_he in FPN and better AVLT performance among healthy older adults. A little to our surprise, we found no difference in the lateralization pattern of FPN between the healthy group and the VCIND groups. However, the significant correlations between LI_he of FPN and AVLT scores in healthy older adults were absent in patients with subcortical VCIND. These findings suggest that subcortical VCIND does not completely disrupt the lateralization pattern of the FPN, but instead selectively impairs the functioning of the right-lateralized FPN in maintaining normal episodic memory.

### Cognitive training reorganizes functional lateralization of FPN in subcortical VCIND patients

Cognitive training has been proven to be an effective intervention for improving the cognitive functions of patients with subcortical VCIND ^6^. To the best of our knowledge, while there is an increasing body of evidence demonstrating the effect of cognitive training in mitigating aging-related dysfunction of higher cognitive networks ^56–58^, only one study has reported an increased FC between the dorsolateral prefrontal cortex and medial prefrontal cortex following short-term cognitive training in subcortical VCIND patients ^6^. By now, the underlying brain mechanism and timing of changes after training are still unclear. Here we used functional lateralization, a fundamental intrinsic organization characteristic of the brain, to explore how cognitive training influences the functional lateralization of FPN and its association with cognitive functions in patients with subcortical VCIND. We found a significant reduction of interhemispheric FC lateralization in FPN, shifting from right-lateralized to bilaterally symmetric, only in the VCIND training group immediately after the 7-week cognitive intervention. Our training paradigms target multidomain cognition supported by FPN, including flexibility, calculation, problem solving, long-term memory, working memory, processing speed, and attention. In contrast, the VCIND control group, which only received attention and processing speed tasks, showed no significant change in FPN lateralization. Notably, changes in FPN lateralization were only observed in the VCIND training group at the end of the 7-week intervention, but not at the 6-month follow-up. This is consistent with previous findings in subcortical VCIND patients that a significant improvement in MoCA and strengthened internetwork functional connectivity after the 7-week cognitive training disappeared at the 6-month follow-up, suggesting that a 7-week training period may have been too short to yield long-term effects ^6^.

Moreover, consistent with the compensation hypothesis, the alterations in FPN lateralization after intervention did contribute to better task performance in the TMT (TMT B and TMT B-A) as well as AVLT (immediate recall and delayed recall) in the VCIND training group. Specifically, the greater changes towards bilateral or even left-lateralized in FPN after 7-week cognitive training, the better executive and memory functions. As executive dysfunction and verbal memory deficit are two prominent impairments associated with subcortical VCIND ^28,29^, our results provided convincing and novel evidence that cognitive training could alleviate executive dysfunction and memory impairment in those patients. This is achieved by rebalancing the interhemispheric functional lateralization of FPN, which allows for compensation by reorganization from neural resources in the left hemisphere. Increases in activity or functional connectivity in the contralateral hemisphere, accompanied by lateralization changes in older adults compared to young adults or following a pathological insult that leaves behavioral functions intact, are frequently attributed to neural compensation ^9,10,19^. Compensation may occur in older adults when they use a neural mechanism that is not available to younger individuals to respond to aging-induced losses ^59^. Similarly, in patients with stroke, a focal ischaemic insult often results in the extensive recruitment of unaffected, remote brain areas ^60–62^. In general, the extent of focal neural damage and the severity of behavioral impairment correlate with compensatory recruitment ^61^, altered functional connectivity ^62^, and greater functional reorganization ^63^ in remote areas. Thus, the significant correlation between a more bilateral or left-lateralized FPN and successful executive and memory performance in the current study is in line with previous findings in recovery from stroke, as well as the HAROLD ^11^ and the STAC-revised models in aging research ^48^, and extend these theories of compensation by reorganization to subcortical VCIND patients following cognitive training. Furthermore, the absence of brain-behavior correlation at the 6-month follow-up indicates that continuous cognitive training is highly recommended for patients with subcortical VCIND to sustain cognitive improvements.

The present study is subject to several limitations. First, a longer intervention period is needed to observe the sustainable benefits of cognitive training and clarify its underlying brain mechanism. Second, although this study used well-defined inclusion and exclusion criteria for subcortical VCIND, we can’t fully rule out the possibility of mixed pathologies, such as concomitant AD. We excluded individuals with indications of atrophy of the hippocampus or entorhinal cortex, which likely excluded individuals with advanced AD pathology; however, prodromal stages with increased amyloid load could not be excluded. Future studies using AD biomarkers are needed to rigorously evaluate the efficacy of cognitive training in patients with subcortical VCIND and to test whether different pathologies respond differentially to cognitive training. Third, it is recommended to include larger sample sizes of patients and well-matched healthy older adults to validate and complement our findings in the future. Last, it should be noted that we retested the neuropsychological assessments in a short interval without a sufficient washout period, which might induce a learning effect and serve as a confounding factor. In an effort to minimize this learning effect on between-group differences, we employed the mixed-effect model to examine the group × time interaction.

## Conclusion

In summary, from a previously unidentified perspective of resting-state functional lateralization, we found that 1) patients with subcortical VCIND exhibited a right-lateralized FPN similar to healthy older adults, but this right lateralization pattern did not support memory function as it did in healthy older adults; 2) via compensation by reorganizing the functional lateralization of FPN from right-lateralized to bilaterally symmetric, the computerized, adaptive, multidomain cognitive training was beneficial to executive and memory functions in patients with subcortical VCIND after intervention. Thus, healthy older adults and patients with subcortical VCIND may rely on different coping strategies and distinct functional lateralization patterns of FPN to achieve better executive and memory performance. This helps deepen the understanding of the influence of subcortical VCIND on the brain and inspires cognitive training intervention. The correlations between the functional lateralization of FPN and cognitive abilities make it a potential biomarker for evaluating the intervention outcomes and underlying brain plasticity. Although the efficacy and good safety profile of computerized cognitive training recommend its application in patients with subcortical VCIND, more clinical trials are needed for further evidence.

## Data Availability

All data produced in the present study are available upon reasonable request to the authors

## Data availability

The datasets used and/or analysed during the current study are available from the corresponding author on reasonable request.

## Acknowledgments

We would like to extend a sincere thank you to everyone who participated in the study.

